# Time for Action: Assessment of Knowledge on Hazard Analysis and Critical Control Points of Food Handlers in Standard Hotels in Lagos State, Nigeria

**DOI:** 10.1101/2022.02.13.22270814

**Authors:** Habeeb Modupe Lateefat, Sawyerr Olawale Henry, Solomon Adewoye

## Abstract

According to the World Health Organization (WHO), there are two million reported cases of food poisoning in Nigeria with estimated deaths of 200,000 people from food poisoning and 20,000 deaths from exposure to food pesticides annually – children inclusive. These hotels make provision of letting room, food and refreshment services in restaurant, bars and banqueting rooms, conference/mailing rooms and leisure facilities it was expressed that, threat identified with the production of food items can be minimized to a permissible limit or eliminated through the utilization of HACCP procedure. This study assessed the knowledge of HACCP of food handlers in standard hotels in Lagos State. The study adopted a cross-sectional survey research design and was conducted in Lagos. All the hotels classified by the Nigeria Tourism Development Corporation (NTDC, 2001) as either three, four or five star-rated were targeted. A purposive and stratified sampling technique was used to select the study units who constituted the respondents for the study with a total of 31 hotels involved in the study. A questionnaire was used to get information on knowledge on HACCP and its implementation by 31 food handlers. Data were presented in tables. Findings from this study has revealed that under 20% of the respondents in the various hotels use HACCP has the quality control strategy, less than 50% understand the concept of HACCP. Results also revealed that there is an insignificant relationship between years of service in the food establishment and the knowledge on HACCP at P value (0.05). More results revealed that there is no significant relation between formal training and knowledge on food safety. Results also revealed that there is no significant relationship between knowledge of HACCP by food handlers and their implementation in food preparation with P-value at (0.05). This insignificant relationship between formal training and knowledge on food safety could mean that there may be other factors preventing food handlers from implementing food safety practice, this may include time, cost of carrying out the practices or even personal attitude of the food handlers during food handling. Food handlers should be taught food safety practices practically rather than theoretically.

## 1. Introduction

Food has an important role in society through affecting people’s lifestyles, health, and habits, as well as being an important aspect of socializing [1, 2, 3, 4, 5, 6]. Today, hotels must pay greater attention to food safety, environmental protection, justice, human rights, and everything else connected to the ethical content of economic operations in the producing world [2, 7, 8, 9]. According to the World Health Organization (WHO), there is growing concern about food safety in hotels, which is currently estimated to account for more than two million reported cases of food poisoning in Nigeria, with 200,000 people dying from food poisoning and 20,000 dying from pesticide exposure each year – including children. Food poisoning in Nigeria is likely to cost millions of lives and trillions of naira per year come 2050 if suitable steps are not implemented to limit its spread. Food poisoning, unlike certain other health disorders, affects every nation, regardless of income or development level [10, 11, 12, 13, 14, 15, 16, 17, 18, 19]. Several studies have found that food-borne diseases impose a remarkable economic as well as quality-of-life burden on society through acute morbidity and chronic sequelae [2, 5, 6, 20, 21, 22, 23, 24, 25, 26, 27, 28, 29, 30, 31]. Food-borne infections cover a wide range of ailments and are an increasing public and environmental health concern across the world. They are the result of consuming polluted water as well as food, and range from diseases caused by a variety of microbes to those induced by chemical dangers [1, 4, 32, 33, 34, 35, 36, 37, 38, 39, 40, 41]. Thus, while gastro-intestinal symptoms are the most prevalent clinical manifestation of food-borne infections, such diseases can also cause chronic, life-threatening symptoms such as gynecological, or immunological problems, neurological, as well as multi-organ failure, cancer, as well as death. Food-borne illness is caused by a diverse range of bacteria, viruses and parasites, it is present all throughout the planet and causes human sickness almost everywhere [2, 20, 22, 28, 29]. While, staphylococcus food poisoning remains the most frequent food-borne infection in the Global South [22, 29]. The bacteria Campylobacter, Salmonella, and E. coli O157:H7, as well as a group of viruses, cause the most generally recognized food-borne illnesses [22, 29]. Pathogenic strains of Staphylococcus spp. can induce food poisoning due to the heat stable Staphyloccal enterotoxin, which is resistant to gastrointestinal enzymes. Food-borne diseases (E. coli and Salmonella) were shown to be the primary cause of these fatalities. While, food handlers play a crucial role in maintaining food safety throughout the manufacturing, processing, storage, and preparation chains [22, 29]. Contamination by the food handler is responsible for 10 to 20% of food-borne illness outbreaks. Mishandling of food and disdain for sanitary precautions allow bacteria to enter food and, in certain situations, survive and reproduce in sufficient quantities to cause sickness in consumers. Personal hygiene and environmental sanitation are crucial in the spread of food-borne illnesses [2]. Investigations of food-borne illness and its outbreaks throughout the world reveal that, in nearly all cases, they remain caused by a failure to meet acceptable standards in the preparation, processing, and storage of food [2, 31, 42, 43, 44].

There was a report of 60 cases and 3 deaths due to food borne disease with a symptomatic gastro intestinal disorders among people who ate in a funeral service [45]. The deaths were linked to food contamination during processing, preservation and service [46]. Melese *et al*., [47], Tuncer and Akoğlu [75] and Scott-Halsell *et al*., [76] characterizes a hotel as a wide range of property types from bigger units having up to or possibly more than 1000 letting rooms, to littler units perhaps having as not many as 10 or even less. Hotels ordinarily provides various services within the same or of different structure which are regularly accessible for the utilization of the both resident and non-resident of the hotel. These include the provision of letting room, food and refreshment services in restaurant, bars and banqueting rooms, conference/mailing rooms and leisure facilities Hazard analysis Critical Control Points (HACCP) has turned out to be recognized universally as the best methods for guaranteeing food safety. In 2004, the European Union (EU) embraced a few new directions on the hygiene level of food, including one (852/2004/EC) authorizing that operative in 2006, all food vendors actualize techniques dependent on the HACCP standards. Government specialists over the globe, including Canada, Australia and Japan, have embraced or are receiving the HACCP-based food safety wellbeing control framework [48; 49]. According to Scott and Stevenson, [48] it was expressed that, threat identified with the production of food items can be minimized to a permissible limit or eliminated through the utilization of HACCP procedure. There is a need to assess the knowledge of HACCP of food handlers in standard hotels in Lagos State. Thus, this study aimed to assess the knowledge of HACCP of food handlers in standard hotels in Lagos State and its environs. The specific objectives are to evaluate the knowledge of food safety practices of the staff and management as regards HACCP compliance in three, four and five star hotels, assess the level of professional training held by both three four and five star hotel operators and their food-handlers. The study focuses on the following Hypothesis Testing (HT):

Ho_1_: There is no relationship between years of service and understanding of HACCP in three, four and five star hotels in Kano Metropolis.

Ho_2_: There is no relationship between formal training classes and application of quality control strategy in food in three, four and five star hotels in Kano Metropolis.

Ho_3_: There is no relationship between knowledge on HACCP and support of the hotels with implementation of HACCP procedure in three, four and five star hotels in Kano Metropolis.

## 2. Materials and Methods

### Study Design

The study adopted a cross-sectional survey research design and was conducted in Lagos. All the hotels classified by the Nigeria Tourism Development Corporation (NTDC, 2001) as either three, four or five star-rated were targeted. A purposive and stratified sampling technique was used to select the study units who constituted the respondents for the study.

### Study Area [Lagos]

Lagos, located within latitude 6^°^27’14.65” N and longitude 3^°^23’40.81” E in the South-Western part of Nigeria is the largest city on the African continent. Based on the 2006 National Census, Lagos State has a population of 9,013,534. However, according to the New York Times publication of 7 January 2014, Lagos’s population is reported to be over 21 million, comprising of a very diverse population due to heavy migration from other parts of Nigeria and surrounding countries. The Yoruba are the dominant ethnic group. Lagos is Nigeria’s major commercial centre and the megacity would be the fifth biggest economy in Africa if it were a country [50, 51].

**Figure 1:**
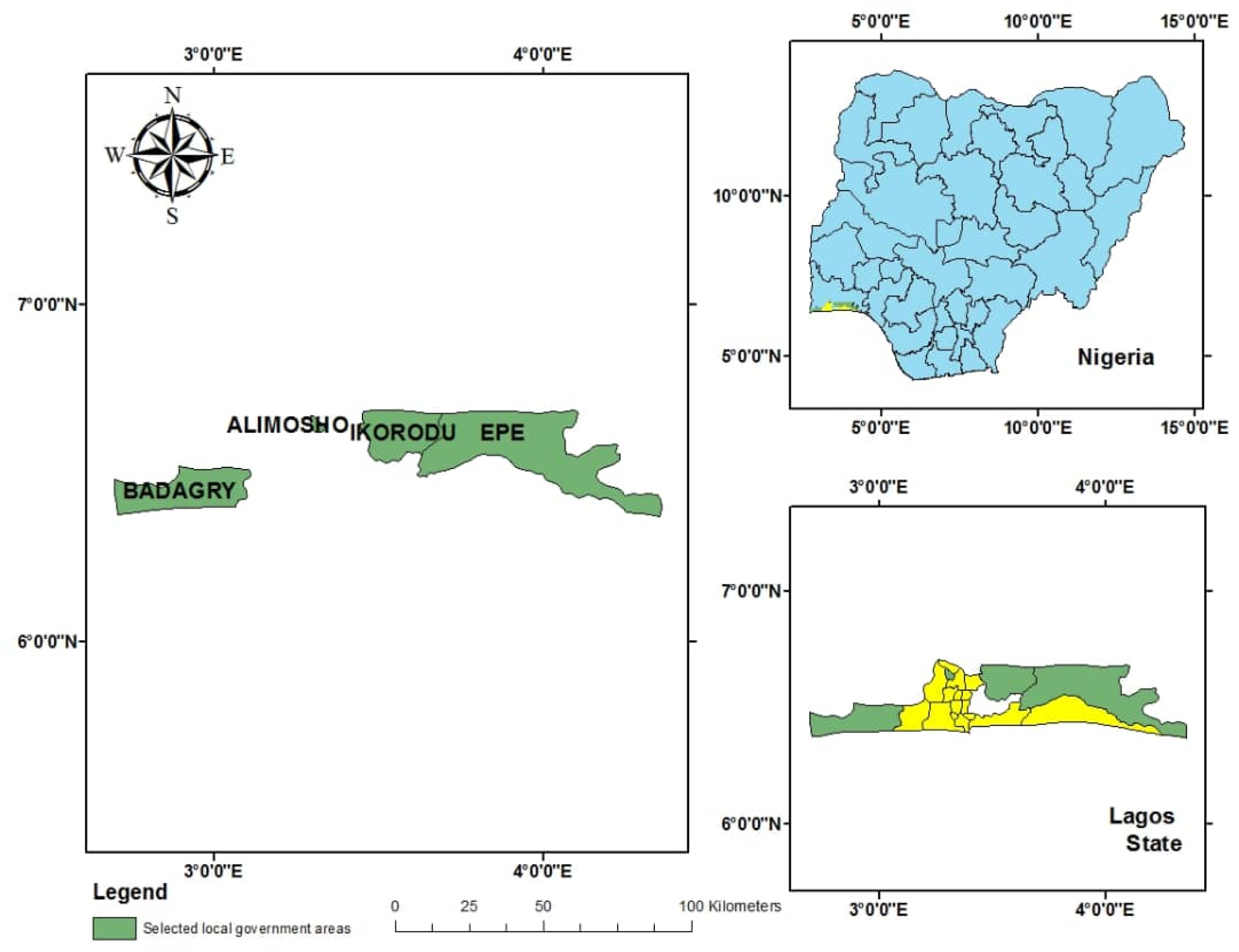
Map of the Study Area in Lagos State.

### Sampling Technique

A purposive and stratified sampling technique was used to select the study units who constituted the respondents for the study with a total of 40 hotels sampled for the study but only 31 hotels allowed entry.

### Study Population

The study population comprised of all food handlers working in the hotel establishments in the study areas.

### Inclusion Criteria

All food handlers in hotels establishments, EHOs at the public health department were included in the study. This survey zeroed in on 3– star to 5-star hotels, that is, hotels with at least 30 rooms, private or public. This classification represents over 70% of the complete hotel stock as per Standard for National Classification and Grading of Hotels and other Serviced Accommodation in Nigeria (2001) created by Nigerian Tourism Development Corporation (NTDC) [74] as a team with Standard Organization of Nigeria (SON).\

### Exclusion Criteria

Food handlers working in other food establishment such as restaurants and street food vendors were excluded from this study.

### Research Instruments

The instruments used for data collections was semi-structured self-administered questionnaire and observation checklist, Sterile container food samples collection and sterile water container.

### Questionnaires

This study considered 31 semi-structured questionnaires administered to food handlers in fast food establishments and government enforcement agents responsible for food premises inspection in the study areas. The questionnaires for the food handlers were divided into sections; A, B, C, D, Close-ended questions were used in both questionnaires.

### Statistical Analysis

The data was cross checked before entry into a computer package called Statistical Package for the Social Sciences (SPSS) version 23. The data was analyzed using techniques such as percentages, figures, tables, as tools of descriptive data analysis which determines the group characteristics with chi-square test.

### Limitations of the study

- Most of the hotels did not permit more than 1 food handlers to be involved in the study because of the time factor.
- Some hotels did not allow entry into their hotels as they feel they are going to sanctioned after the study.

## 3. Results

## 4. Discussions

From the table 1 above, a large proportion of the food handlers were between the ages on 34-41years (62.5%), while 27.5 % of the food handlers were between the ages 26-33years while (10%) were with 18-25 years. This study is relating with research done by Ulusoy and Colakoğlu [52] were most participants were aged 19–40 and over 40 (75.8% and 19.4%, respectively) in the survey on determination of HACCP knowledge of food handlers in Istanbul food businesses. This is similar to studies in Ghana by Annor and Baiden [53] who showed that majority of respondents were under thirty years. At this age, respondents are very young and are able to manage any work given to them. From the table 1 above, (50%) of the food handlers were males and 50% were females. This study is contrasting with a study by Habiballah *et al*. [54] which revealed that women, who generally prepared food, do not form the highest number in restaurant operations. In contrast to the domestic setting where more women are cooks, both men are women are formed as the cooks/chefs in Nigeria hotels as indicated in this study, with more men engaging in culinary activities which used to be a woman’s job. This study is also negating a study by Martins *et al*. [55], where of the 101 food handlers questioned in this study, 96.0% were women. All male participants were in management positions in the “Food handlers’ knowledge on food hygiene: The case of a catering company in Portugal”. From the table 1 above, majority of the handlers in Lagos state revealed that (50%) have OND/NCE qualification while 10% has a qualification in BSc./Msc., while 20% of the food handlers have secondary school qualification, with another (20%) in primary school. This is in contrast with Mukhtar *et al*., [56] where among the 350 respondents, 5.1% of the respondents did not have any formal education, 16.9% have primary education, 71.1% have secondary education, 4.6% have diploma and 2.3% have degree. Previous research found that food safety training increased knowledge regarding food safety issues [1, 2, 3, 4, 44, 57, 58]. From the table 1, a great number of the respondents in this study were Head cooks (55%), (27.5%) were managers, while 17% were cooks. This is dissimilar to the work by Rahman *et al*. [59] with respondents’ position, Cook (23%), Manager (53%). Preparation (23%). In Table 1, when asked about the years of service in the establishment, Majority (92.5%) of them has worked for more than three months in the hotel, when asked the number of training attended, (35%) never attended any training program, (10%) has attended two training programs, (5%) has attended two, (25%) attended three training program, while (7.5%) have attended four training programs (17%) has attended five and above training programs with (62.5%) of the respondents that has attended formal food training programs and (37.5%) of the respondents not attended any formal food training programs. Leaving (67.5%) of the respondents certified and (32.5%) of the respondents not certified. This study is in contrast with Pepple, [60] where all the respondents were certified caterers who had completed course in Food Hygiene in the “Environment and Food Poisoning: Food Safety Knowledge and Practice among Food Vendors in Garki, Abuja – Nigeria”. In a related study [61] also found that less than half of the food vendors studied in Benin City, Nigeria had undergone training in food hygiene and safety. This is also in contrast with the findings obtained from the study of Bas *et al*. [62] indicated that 28.4% of managerial staff and 56.3% of basic food handlers had not received basic food hygiene training. In a study, 55% of the 444 food handlers received formal food hygiene training, and 63% of managers had undertaken formal food hygiene training in UK food businesses [63]. Results from this study implies that a large number of the respondents have undergone formal food training sessions and have been certified, thus may have better knowledge of food safety practices in food production,

**Table 1:** Demographic Characteristics.

|  | *n* | *%* |
| --- | --- | --- |
| ***Age*** |  |  |
| *18-25 yrs* | 11 | 27.5 |
| *26-33yrs* | 25 | 62.5 |
| *34-41yrs* | 4 | 10.0 |
| *50yrs and above* | 0 | 0.0 |
| ***Gender*** |  |  |
| *Male* | 19 | 47.5 |
| *Female* | 21 | 52.5 |
| ***Education*** |  |  |
| *Primary* | 0 | 0.0 |
| *Secondary* | 0 | 0.0 |
| *OND/NCE* | 20 | 50.0 |
| *BSC/B.ED/HND* | 20 | 50.0 |
| ***Position*** |  |  |
| *Manager* | 11 | 27.5 |
| *Head cook* | 22 | 55.0 |
| *Cook* | 7 | 17.0 |
| *Waiter* | 0 | 0.0 |
| *Cleaner* | 0 | 0.0 |
| ***Years of service*** |  |  |
| *2 months* | 3 | 7.5 |
| *Above 3months* | 37 | 92.5 |
| ***No. of_Training Program attended*** |  |  |
| *none* | 14 | 35.0 |
| *one* | 4 | 10.0 |
| *two* | 2 | 5.0 |
| *three* | 10 | 25.0 |
| *four* | 3 | 7.5 |
| *five & above* | 7 | 17.5 |
| ***Formal_training_catering_classes*** | 25 | 62.5 |
| *Yes* |  |  |
| *No* | 15 | 37.5 |
| ***Certificate training*** |  |  |
| *Yes* | 27 | 67.5 |
| *No* | 13 | 32.5 |
| ***How_often_do_you_go_for_training*** |  |  |
| *Not at all* | 1 | 2.5 |
| *Quarterly* | 7 | 17.5 |
| *6 months* | 10 | 25.0 |
| *Yearly* | 22 | 55.5 |

### Knowledge on Quality Control

From the table 2 above, the respondents were asked if they know any quality control measures in food establishment, majority of the respondents (80%) said yes while a few (80%) said no in the sampling of selected hotels in Lagos state. When asked the type of quality control measure they use in their establishment. Most of them (37.5%) said Standard Operation Procedure (SOP), (25%) mentioned Good manufacturing practice (GMP), (17.0%) standard sanitation operation procedure (SSOPs), a few (15%%) mentioned Good agricultural practices (GAP) while another (5%) respondents mentioned Hazard analysis and critical control points (HACCP). Findings from this study shows that the respondents have different knowledge on food quality control measures which may ensure safe production of food. In table 2 above, when asked about the where they will apply the quality control measures used, (12.5%) of the respondents said in the kitchen during cooking only, (30%) said when receiving food products only, (0%) respondents mentioned during storage, while majority of the respondents (57.5%) mentioned all of the above. This shows that majority of the respondents understands the principles of food safety that food safety applies to all processing steps in food production. This is in line with the study carried out by Food Safety Authority of Ireland, 2001, 89% and upwards of process steps were said to have a food safety procedure associated with them in the Survey of the Implementation of HACCP (Hazard Analysis and Critical Control Point) and Food Hygiene Training in Irish Food Businesses. This is in contrast with the work by Muinde and Kuri, [64] where majority of the quality control (92%) strategy while receiving products just (8%) was applied in the kitchen only in the “hygiene practices in urban restaurants and challenges to implementing food safety and hazard analysis critical control points (HACCP) programmes in Thika town, Kenya”. In this study, the application of quality control strategy in all food process steps shows a clear understanding of its importance by the food handler. From the table 2 above, it was revealed that they heard of these quality control measures, (80%) of the respondents said they heard from Environmental health officers in the states and local government, (20%) had heard it from NAFDAC, (10%) had heard it from a customer (2.5%) had heard from mass media and others cannot remember where they heard it from. Results from this study, observes that majority of the respondents got their understanding quality control measures from EHOs in their jurisdiction, this implies that EHO’s are saddles with the responsibilities of training of food handlers on different quality control strategies. This study is in conformity with a study by Muinde and Kuri, [64] where majority (92%) of the respondent heard of quality control strategy from public health staffs and a few (8%) heard it from council staff in the “hygiene practices in urban restaurants and challenges to implementing food safety and hazard analysis critical control points (HACCP) programmes in Thika town, Kenya” Results from this study, observes that majority of the respondents got their understanding of quality control measures from EHOs in their jurisdiction, this implies that EHO’s serves a function in training of food handlers on different quality control strategies. This study is in conformity with previous study by [8, 9]. Similarly, in table 2 above, Majority of the respondents (77.5%) said they have not heard of it while (22.5%) said they have heard of the term HACCP. This present study is in contrast with study by Walker *et al*, [63] reported, 42% of managers had heard of HACCP and 65% could not explain what it involved but in conformity with a study by Bas *et al*., [65] declared, approximately 57% of managers they surveyed told that they had heard of the HACCP system, 18.3% of those who had heard of the term said they had a HACCP team, and the minority (21.7%) said that food handling was carried out in accordance with the principles of HACCP system. To achieve the successful implementation of HACCP, the concept must be understood first by the managers of the establishments [66]. After providing that, it is important to remember that the system needs the involvement of all personnel in the HACCP methodology and philosophy. From table 4 above, when asked if they have a written hygiene statement apart from HACCP, majority of the respondents (55%) said they do not have while a few of the respondents (45%) said they have a written hygiene statement. Results from this study interprets that with the majority of hotels without written food hygiene statement, food handlers may not remember to implement food safety practices unlike when it is written and even pasted around the kitchen environment. This study does not conform with the Food Safety Authority of Ireland, 2001 with (53%) of the respondents having a written hygiene statement. Similarly, Worsfold and Griffith [67] reported that many small food businesses lacked written hygiene procedures, performance standards and personal hygiene rules. A written hygiene statement would serve as a reminder for food handlers in keeping regular hygiene practice in food production. From table 2 above, when asked the statement “how would you rate your understanding on HACCP”, a great proportion of the respondents (70%) said “fair”, (5%) said “poor”, (5%) said “very poor”, (20%) said “Good”. This work is also similar to what is observed in the studies from Ireland by Bolton *et al*., [68] and from Brazil by Rebouc as *et al*., [69] observed that nearly 35.0% of the participants knew what the HACCP means. This study does not conform with a study by Rombo and Muoki [70] where majority of the respondent (60%) have understanding of HACCP as a quality control measure in the Hygiene practices in urban restaurants and challenges to implementing food safety and hazard analysis critical control points (HACCP) programmes in Thika town, Kenya. Furthermore, table 2 above, majority of the respondents (60%) said there is no implementation of HACCP in their establishment and they do not have a HACCP team while few (40%) said they implement HACCP in their establishments and they have a HACCP team. This study is in contrast with a study by Doneva-Sapceska and Alchevska [71] where (86%) of the respondents implement HACCP in their establishment and have a HACCP team or an individual with an overall responsibility on HACCP in the “Analysis of effectiveness of HACCP system in small restaurants in Skopje”. Findings from this study was that HACCP has not been widely used and that this had a negative impact on the general food hygiene standards and food-handling practices of personnel. HACCP has yet to become a legal requirement for the Turkey food industry. From table 3, A p-value results shows 0.332 with p-value set at (0.05). The null hypothesis is accepted with a p-value greater than 0.05. It shows that that there is no significant relationship between the years of service in the food establishment and the understanding of HACCP. This relates with the work by Webb and Morancie, (2015) [72], where the correlation analysis revealed there was a weak and indirect association between the length of employment in the foodservice industry and food safety knowledge. From tables 4, A p-value results shows 0.386 respectively with p-value set at (0.05). This revealed that there is no significant relationship between the formal training of food handlers and application quality control strategy in food establishment with a p-value (0.005). The Null hypothesis is accepted with a p-value greater than 0.05. This shows that the formal training of food handlers does not have a positive impact in the application of food safety strategy in different steps in the food establishment in this study. This means there could be other reasons that prevents food handlers who has undergone formal training and implementing a quality control strategy to food, these factors could be time, cost or laziness on the part of the food handlers. In table 5 above, A p-value results shows (0.279) with p-value set at (0.05). which means there is no significant relationship between the Knowledge on HACCP and the implementation in the hotels. This is similar to the study by Ko [73] where there was an insignificant relationship identified between knowledge and HACCP practices indicated that the former does not directly influence the latter, except through the mediating influence of attitude. Results from Kano and Delta revealed that there is a significant relationship between the Knowledge on HACCP and the implementation in the hotels.

**Table 2:** Knowledge on Quality Control.

| *Statements* | *n* | *%* |
| --- | --- | --- |
| ***Do you know any quality control measures for food establishment?*** |  |  |
| *Yes* | **32** | **80** |
| *No* | **8** | **20** |
| ***If yes, which of the measure;*** |  |  |
| *Good Manufacturing procedure* | 6 | 15.0 |
| *Good Agricultural Procedure* | 15 | 37.5 |
| *Standard Operation Procedure* | 10 | 25.0 |
| *Standard Sanitation Operation Procedure* | 7 | 17.5 |
| *Hazard Analysis and Critical Control Points* | 2 | 5.0 |
| ***where would you apply this quality control strategy in your food*** |  |  |
| *Receiving Food Products only* | 12 | 30.0 |
| *Storing food products only* | 0.0 | 0.0 |
| *Kitchen during cooking only* | 5 | 12.5 |
| *All of the above* | 23 | 57.5 |
| ***What kind of quality control measures do you now apart from the mentioned?*** |  |  |
| ***From where did you know these strategies?*** |  |  |
| *Environmental health Officers from the State and Local government* | 32 | 80 |
| *NAFDAC* | 4 | 20.0 |
| *Customer* | 2 | 10.0 |
| *WHO/FOA* | 0 | 0.0 |
| *Mass media* | 1 | 2.5 |
| *Can't Remember* | 1 | 2.5 |
| *Have you heard of the term "Hazard Analysis and Critical Control Points"* |  |  |
| *Yes* | 31 | 77.5 |
| *No* | 9 | 22.5 |
| *Do you have a written food Hygiene statement Apart from HACCP* |  |  |
| *Yes* | 18 | 45.0 |
| *No* | 22 | 55.0 |
| *How would you rate your understanding on HACCP?* |  |  |
| *Excellent* | 0.0 | 0 |
| *Good* | 8 | 20.0 |
| *Fair* | 28 | 70.0 |
| *Poor* | 2 | 5.0 |
| *Very Poor* | 2 | 5.0 |

**Table 3:**
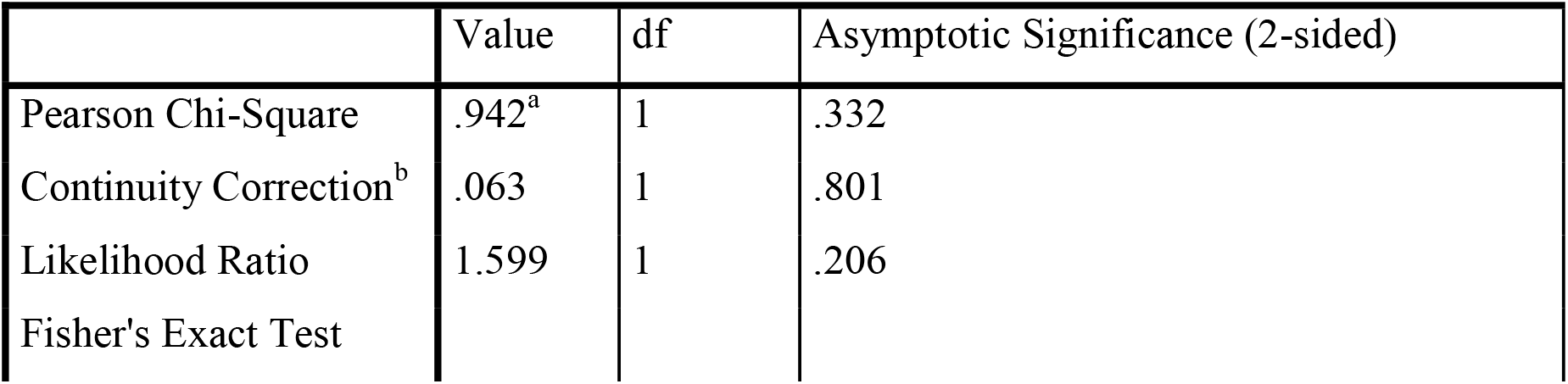
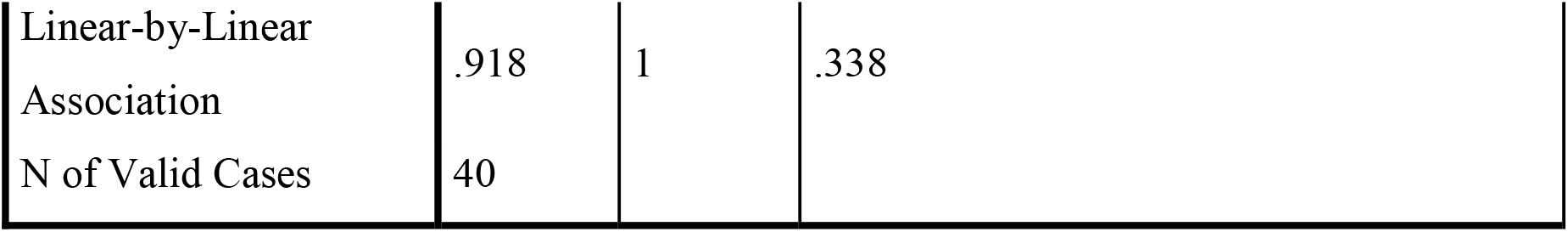
Relationship between years of service and understanding of the term” Hazard Analysis and Critical Control Points.

**Table 4:**
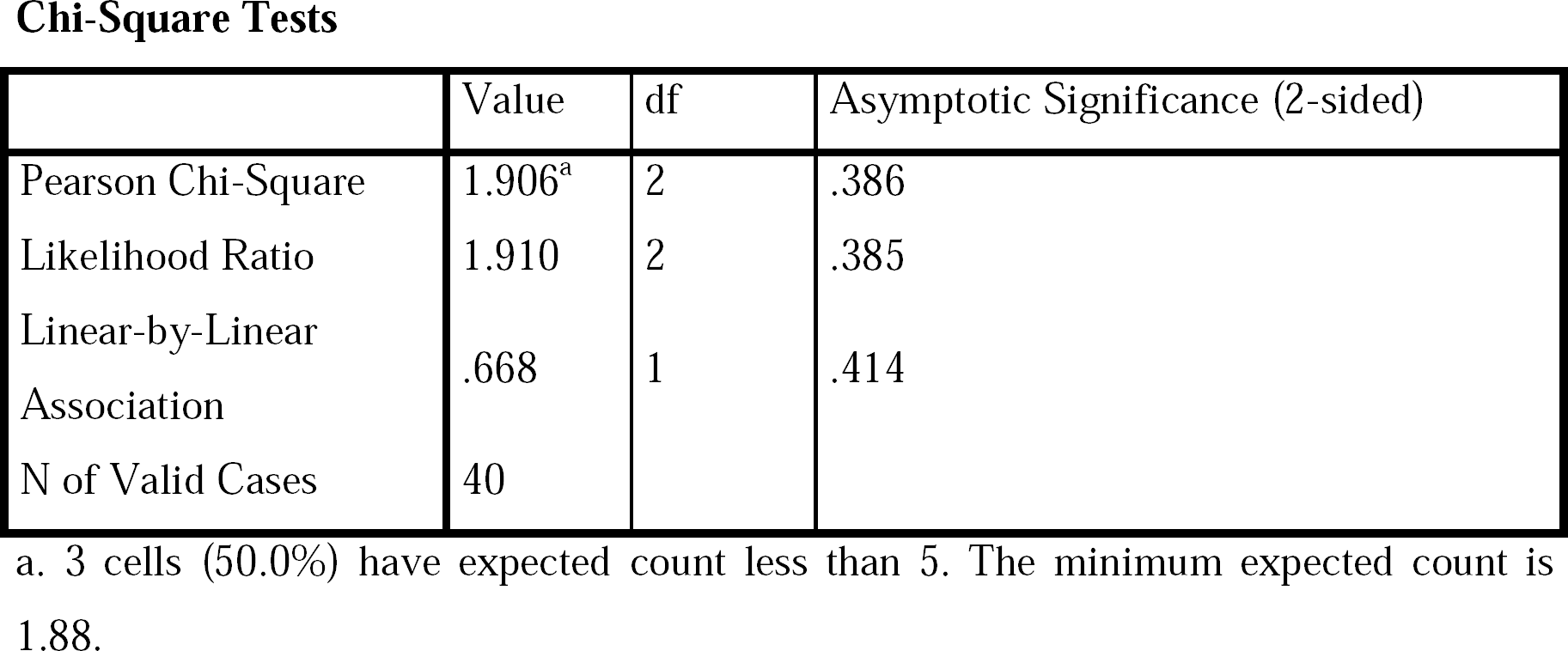
Relationship between Formal Training and Application of Quality Control Strategies in the Food Preparation

**Table 5:** Have you heard of the term” Hazard Analysis and Critical Control Points” * Generally, how supportive or unsupportive is your hotel with implementation of HACCP procedure.

|  | Value | df | Asymptotic Significance (2-sided) |
| --- | --- | --- | --- |
| Pearson Chi-Square | 1.171<sup>a</sup> | 1 | .279 |
| Continuity Correction<sup>b</sup> | .484 | 1 | .487 |
| Likelihood Ratio | 1.151 | 1 | .283 |
| Fisher's Exact Test |  |  |  |
| Linear-by-Linear Association | 1.142 | 1 | .285 |
| N of Valid Cases | 40 |  |  |

## 5. Conclusion

Findings from this study has revealed that under 20% of the respondents in the various hotels uses Hazard Analysis and Critical Control Points (HACCP) has the quality control strategy, less than 50% understand the concept of HACCP. Results also revealed that there is an insignificant relationship between years of service in the food establishment and the knowledge on HACCP at p-value (0.05). This could mean that food handlers may not upgrade, regarding new food safety methods or may choose to stick their old knowledge on food safety. Findings from this study also revealed that there is no significant relation between formal training and knowledge on food safety. This could mean that there may be other factors preventing food handlers from implementing food safety practice, this may include time, cost of carrying out the practices or even personal attitude of the food handlers during food handling. Results also revealed that there is no significant relationship between knowledge of HACCP by food handlers and their implementation in food preparation with p-value at (0.05). This could be as a result of lack of time, extra cost in implementation of HACCP and unavailability of equipment’s. Conclusively, the findings of this study may spur the expansion of information programs aimed at expanding food safety knowledge and raising awareness levels among the various hotels. Furthermore, the investigations allow for the consideration of coordinated measures across the various hotels in order to raise customer and staff knowledge of hotel-related features.

## 6. Recommendations

i. Regular training programmes should be organized for handlers by their establishment by the Environmental Health Officers to acquire new knowledge on food safety.

ii. Food handlers should be taught food safety practices practically rather than theoretical methods.

iii. Hotel operators should employ professional HACCP consultant in the implementation of HACCP systems in their hotels.

iv. Hotels should prepare HACCP plans for different foods in their hotels to allow timely implementation of the system.

v. EHO’s should be equipped with tools for implementation of food safety strategy and HACCP system.

## Data Availability

All data produced in the present work are contained in the manuscript

## Abbreviations

HACCP: Hazard analysis critical control point
WHO: World health organization

## Data Availability

Data used to support the findings of this study are included within the article.

## Acknowledgements

Authors are thankful for the hotel kitchen staff who participated in this research study and also Sanitarian Raimi Morufu Olalekan for editing the article. This study was a part of a PhD Thesis of Habeeb Modupe Lateefat, (Department of Environmental Health Science, Kwara State University, Malete).

